# Instability as Signal: Diagnosing Latent Heterogeneity in Multimodal Survival Models

**DOI:** 10.64898/2026.10.13.26365414

**Authors:** Rishika Ray, Sayar Singh Shekhawat

## Abstract

Multimodal deep learning models for cancer survival prediction frequently exhibit unstable and irreproducible performance, limiting their clinical applicability. Rather than proposing a new predictive architecture, this work presents a diagnostic investigation into the sources of this instability. Using a transformer-based deep fusion model combining RNA sequencing and histopathology data from the TCGA-BRCA cohort as a diagnostic probe, we demonstrate that global survival prediction is highly unstable across data splits. We then analyze the learned multimodal embeddings in an unsupervised manner, revealing stable latent patient subgroups with significantly divergent survival outcomes. These subgroups are not explained by canonical receptor status or immune infiltration, but instead align with tumor-intrinsic proliferative activity. Conditioning survival modeling on the largest discovered subgroup recovers strong and stable prognostic performance, substantially outperforming unimodal baselines. These findings indicate that survival predictability is conditional on latent heterogeneity and suggest that heterogeneity-aware modeling strategies are essential for robust multimodal prognostication. This work establishes a methodological framework for interpreting instability in multimodal survival learning.

## I. Introduction

The estimation of a patient’s future clinical outcome, such as survival time, disease progression, or recurrence risk, based on biological, pathological, and clinical data is known as cancer prognostication. Since the realisation that no single data modality can adequately capture the complexity of tumour biology, multimodal deep learning has emerged as a potential paradigm. RNA sequencing provides transcriptional programs, oncogenic signalling pathways, and cellular states. On the other hand, histopathology images encode spatial organisation, morphological patterns, and microenvironmental context. Integrating these two modalities of data should produce more accurate survival predictions as compared to unimodal methods [1], [2].This idea was tested out on pancancer settings and increasingly sophisticated fusion strategies [2], [3]. More recently, transformer-based attention-based architectures have been suggested for the construction of complex cross-modal interactions, thereby expanding the representational capacity of multimodal models [4]. These efforts suggest that deep multimodal fusion can uncover prognostically relevant signals that are inaccessible to individual modalities.

The translation of multimodal survival models into reliable clinical tools remains limited in spite of the progress. A persistent and under-discussed problem is the instability of model performance shown by substantial variance of reported concordance indices across random data splits, cross-validation folds, or minor changes in preprocessing [5]. Although models achieve competitive average performance, they behave almost randomly in some validation sets. This undermines the confidence in their predictions [6]. This phenomenon is particularly seen in survival analysis, where the datasets are generally small, event rates are low, and the model outputs affect high clinical decisions [7].

This performance instability is reduced through refinement of architecture, hyperparameter tuning or increased regularisation, and is implicitly treated like a technical nuisance in most of the existing studies. This perspective assumes the existence of a single, global prognostic function for the entire patient cohort and that the cause of instability is mainly from insufficient modelling capacity or suboptimal training procedures. This paper explores the possibility that this assumption is flawed for heterogeneous diseases like cancer. Instability may be a diagnostic signal that shows the presence of unmodelled patient heterogeneity [8].

Breast cancer is a very good example of this challenge. It is known that breast cancer is not a single disease entity; it is a collection of biologically distinct subtypes with divergent molecular characteristics and clinical trajectories [9]. These differences are present at the receptor level or gene expression signatures as well as in tumour morphology and treatment response. This implies that patients within a single cohort may follow fundamentally different survival-generating mechanisms. A global survival model trained on such a diverse population will learn an “average” representation. It may fit no subgroup well, therefore, resulting in high variance in performance across data splits.

This issue is aggravated in multimodal settings. Multimodal fusion increases representational richness as well as model capacity and the effective degrees of freedom. This imbalance between model complexity and available outcome information in event-limited survival datasets like the TCGA (The Cancer Genome Atlas) can increase variance and overfitting [7]. But this does not imply that multimodal models are unsuitable for survival prediction. It suggests that their failure modes are more subtle and closely tied to the structure of the underlying population than is accepted.

A related issue is the evaluation of survival models. The most common used metric for assessment of survival performance is the concordance index (C-index) [10]. Though the C-index is an important summary statistic, it collapses model behaviour across all patients into a single scalar value. In heterogeneous cohorts, this aggregation can conceal strong subgroup-specific performance and hide systemic failure modes. As pointed out earlier, reliance on global Cindex values may lead to deceptive conclusions about model utility, in particular when performance varies substantially across patient subpopulations [5]. As a result, instability in global metrics should be used as a potential indicator of latent structure rather than dismissed as noise.

Recent studies of machine learning in medical imaging are focusing on the importance of understanding why models fail [6]. These critiques argue that most of the reported advances are weakened by inadequate evaluation practices, hidden dataset biases and a lack of diagnostic analysis. In the context of multimodal survival prediction, there is a need to move beyond architecture-centric benchmarks and toward investigations into the relationship between model behaviour and underlying biological heterogeneity.

This study adopts a diagnostic perspective. This paper deals with the investigation of the causes of instability in a standard transformer-based multimodal survival model, instead of proposing a novel fusion architecture or optimising for maximum global C-index. We hypothesise that performance instability is not noise but a structured signal arising from latent patient heterogeneity. Deep multimodal fusion can expose this heterogeneity even when it fails to produce stable global predictions.

To test this theory, we conduct a sequential empirical investigation using the TCGA Breast Invasive Carcinoma (TCGA-BRCA) cohort [11]. First, the global multimodal survival prediction is found, and its instability is explicitly characterised. Then, the trained model is repurposed as a representation learning tool. Fused multimodal embeddings are extracted, and unsupervised analysis is performed. This helps to assess whether prognostically relevant structure emerges in the learned interaction space or not. It should be noted that this analysis does not use survival outcomes, receptor status or clinical labels.

The multimodal embeddings divide patients into stable subgroups with markedly different survival outcomes. These subgroups are not recoverable from unimodal representations or naive feature concatenation. This shows that deep multimodal fusion captures interaction-level information that is not visible to individual modalities. Also, the discovered subgroups cannot be explained by canonical breast cancer markers such as receptor status. Instead, it can be aligned with tumour-intrinsic biological characteristics, particularly proliferative activity [9].

Survival prediction becomes accurate and stable when conditioned on a homogeneous patient subgroup. Here, multimodal modelling will substantially outperform unimodal baselines. It is consistent with histopathology and molecular data that provide complementary prognostic information [1], [3]. Across resampling, conditional modelling exhibits minimal variance. Negative control experiments confirm that instead of statistical artefacts, these gains reflect genuine prognostic signal [6].

Together, these findings support a reframing of multimodal survival modelling. Instead of asking whether optimising global state-of-the-art performance on heterogeneous cohorts is sufficient [6], the more fundamental question is how multimodal representations encode latent biological structure. This structure conditions survival predictability [8], [9]. By shifting the focus from aggregate performance metrics toward diagnostic analysis [5], this work highlights the importance of understanding when and for whom multimodal survival prediction is feasible. This also motivates heterogeneity-aware modelling strategies as a path toward more robust and clinically meaningful prognostication.

## II. Related work

### A. Multimodal Survival Prediction

An active area of research is multimodal survival prediction. It has been motivated by the hypothesis that integrating heterogeneous biomedical data sources can yield more accurate and biologically grounded prognostic models than unimodal approaches. Early work by Mobadersany et al. [1] and subsequent pan-cancer analysis by Kather et al. [12] showed that combining histopathology images with genomic features using convolutional neural networks improved outcome prediction compared to either modality alone. This study provided one of the first large-scale demonstrations that complementary prognostic information is contained in molecular profiles and histological morphology.

Subsequent work extended multimodal survival modelling to pancancer settings. Cheerla and Gevaert [2] proposed a deep learning framework integrating genomics, clinical variables, and imaging-derived features across multiple cancer types. Their results showed improved average concordance indices compared to unimodal baselines. This reinforces the potential of multimodal learning. Vale-Silva and Rohr [3] also explored multimodal architectures for long-term cancer survival prediction. They showed the benefits of feature fusion in deep neural networks.

In spite of these promising results, most multimodal survival studies focus on improving global predictive performance. Model evaluation is done using mean C index values across cross-validation folds, with limited discussion of performance variance or subgroup-specific behaviour [6]. Therefore, the robustness and interpretability of these models in heterogeneous clinical populations remain open questions. Furthermore, reliance on global concordance indices can be misleading, as these metrics may be biased by the censoring distribution and fail to capture time-dependent performance variations [5].

### B. Deep Learning Models for Survival Analysis

DeepSurv [13] introduced a neural network formulation of the Cox proportional hazards model, enabling nonlinear risk modelling while preserving the interpretability of hazard-based survival analysis. DeepHit [14] proposed a discrete-time formulation capable of modelling competing risks.

Studies of deep learning for survival analysis highlight the diversity of proposed architectures. It includes recurrent networks, multitask learning, and attention-based models [7]. These methods differ in formulation and optimisation objectives. But they all want to improve global performance metrics. Importantly, few studies interrogate the stability of these metrics across heterogeneous patient populations or explore whether performance variability reflects underlying biological structure [6], [8].

### C. Transformer-Based Fusion

In medical applications for multimodal learning, attention mechanisms and transformer architectures have become very popular. Through self-attention and cross-attention mechanisms, transformers are a flexible framework for modelling interactions within and between modalities. Multi-modal survival models leverage these capabilities to capture conditional relationships between histopathology, genomics, and other data sources.

The PS3 framework [4] is an example. It integrates pathology reports, histology images, and biological pathways using a multimodal transformer. PS3 demonstrates improved survival prediction performance across multiple cancer cohorts by modelling interactions between modalities. Evaluation primarily centres on global performance metrics, leaving open questions about how learned representations behave in heterogeneous populations, just like earlier approaches.

### D. Image-Based Genomic Signal Discovery

Several studies have shown that deep learning models can extract molecular information directly from histopathology images. Kather et al. [12] showed that convolutional neural networks trained on WSIs (Whole Slide Images) could predict clinically actionable genetic alterations across multiple cancer types. These provide strong evidence that morphological patterns encode latent molecular signals.

Multimodal frameworks like MOMA [15] also emphasise explainability and cross-cohort validation in integrating pathology images with multi-omics data. These studies focus on classification or molecular inference rather than survival modelling. They also establish a precedent for deep learning models uncovering non-trivial biological structure through multimodal integration.

### E. Patient Heterogeneity and Evaluation Pitfalls

Biological heterogeneity is a defining feature of cancer. Marusyk and Polyak [8] expressed the evolutionary and microenvironmental origins of tumour heterogeneity. They highlighted its implications for treatment resistance and clinical outcomes. Large-scale molecular analyses, like those conducted by Hoadley et al. [9], also demonstrate that tumours cluster into biologically distinct subtypes across tissues of origin.

From the context of machine learning, heterogeneity presents a challenge for global predictive modelling. Unsupervised clustering has been used to identify latent structure in high-dimensional biological data for a long time, but clustering stability and interpretability remain critical concerns. Von Luxburg [16], [17] provides a theoretical foundation for clustering methods and stability assessment. She shows the importance of robustness to initialisation and sampling variability. Analogous challenges arise in single-cell transcriptomics, where clustering results can vary depending on algorithmic choices and preprocessing [18]. These observations need careful validation when interpreting unsupervised subgroup discovery, particularly in clinical contexts.

### F. Positioning of the Present Study

Few multimodal survival studies investigate why models fail or how instability relates to underlying biological structure [7]. This gap motivates the present work, which positions instability as a signal pointing to latent heterogeneity. This study adopts a diagnostic framework to understand the relationship between multimodal fusion, heterogeneity, and survival predictability. By analysing learned multimodal embeddings and conditioning survival modelling on discovered subgroups, this work bridges multimodal representation learning, unsupervised subgroup discovery, and survival analysis in a unified framework.

## II. Methods

### A. Study Design and Data Description

This study follows a diagnostic methodological framework that aims to understand the causes of instability in multimodal survival models. Here, performance variance is treated as noise and an investigation is done to determine whether it reflects latent biological heterogeneity within the patient population. The analysis has three sequential stages:

1. Construction and evaluation of a global multimodal survival model
2. Unsupervised analysis of learned multimodal representations to identify latent patient subgroups,
3. Conditional survival modeling within discovered sub-groups

This approach is motivated by previous work that has shown that aggregate evaluation metrics may mask systematic failure modes in heterogeneous clinical datasets [5], [6].

All experiments were performed on the TCGA-BRCA dataset [11], which consists of 1020 patients with matched RNA sequencing and histopathological data. Survival outcomes were specified based on overall survival time and censoring variables. A total of 145 death events were recorded,which places the study in an event-limited setting where the effective capacity of models is limited by the number of observed events, rather than the size of the dataset itself [10]. This setting informed all architectural and methodological decisions. Figure 1 shows a high-level diagram of the complete analysis pipeline.

**Fig 1.**
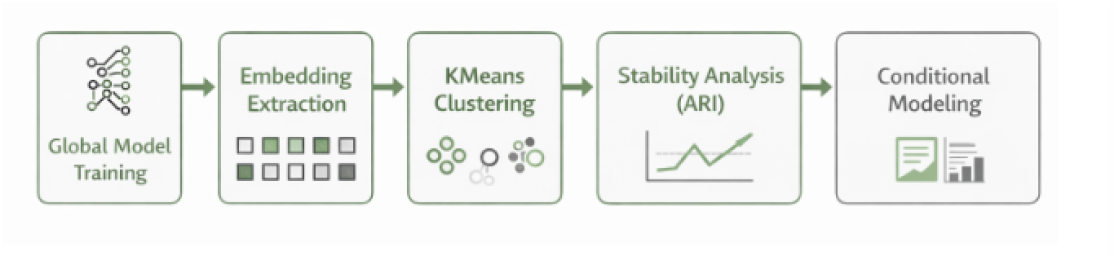
Overview of the diagnostic multimodal survival modeling pipeline. The framework progresses from global modeling to latent structure discovery and conditional evaluation.

### B. Multimodal Data Representation and Model Architecture

The RNA sequencing data was represented using low-dimensional embeddings derived through principal component analysis (PCA) on normalised gene expression matrices. Raw RNA-seq data generally consists of tens of thousands of gene features, which is significantly higher than the number of available samples in the TCGA-BRCA cohort (N = 1020). Using such high-dimensional input would lead to extreme overfitting and poor generalisaton, that is, the model would be unable to learn. Thus, PCA was used to convert gene expression profiles into a lower-dimensional space. It retained the dominant axes of transcriptional variability and also suppressed noise and redundant features. Each patient was shown using a 128-dimensional vector embedding, which is a compact and informative representation of the molecular state. For numerical stability, more preprocessing was done. Values that are invalid, like NaNs, were replaced with zeros to prevent propagation of undefined values during optimization. Feature values were clipped to a fixed range to reduce the influence of extreme outliers. Extreme outliers lead to unstable gradients in deep neural networks. These steps collectively ensure stable and reliable integration of RNA-seq data into the multimodal model.

The histopathology whole-slide images WSIs were extremely high resolution; thus, direct end-to-end processing is computational infeasible. Due to this, each slide was first partitioned into smaller image patches following foreground segmentation to exclude background regions and retain tissue-containing areas. Each patch was then encoded into a fixed-length feature vector using a pretrained CNN. This approach extracts high-level morphological features that capture relevant histological patterns such as cellular architecture, tissue organisation, and microenvironmental context. The resulting 2048-dimensional feature vectors provide a compact and semantically meaningful representation of each patch. This patch-level representation enables efficient modeling while preserving local morphological variability across the slide. By representing WSIs as collections of feature vectors rather than raw images, the approach significantly reduces computational complexity and facilitates downstream integration with multimodal learning frameworks. Similar feature extraction strategies have been widely adopted in prior multimodal and computational pathology studies [2], [15].

To generate patient-level image features, a fixed number of *K* = 8 patches were randomly sampled from available tissue areas. This selection balances spatial heterogeneity retention and model complexity within the event-limited setting. Although larger values of K may retain more morphological information, they increase the degrees of freedom and the risk of overfitting. Random sampling was deliberately chosen to prevent selection bias and survival information on patch selection, allowing for evaluation under reduced assumptions. The multimodal model architecture is a transformer-based framework that combines RNA and image data using cross-attention. A diagram of the architecture is provided in Figure 2. Both modalities are first projected into a common latent space via linear layers. The sequence of image tokens is then processed by self-attention to model relationships between spatially separate regions, such as tumour-stroma interfaces or heterogeneous tumour areas. The resulting token embeddings are then aggregated into a single image embedding.

**Fig 2.**
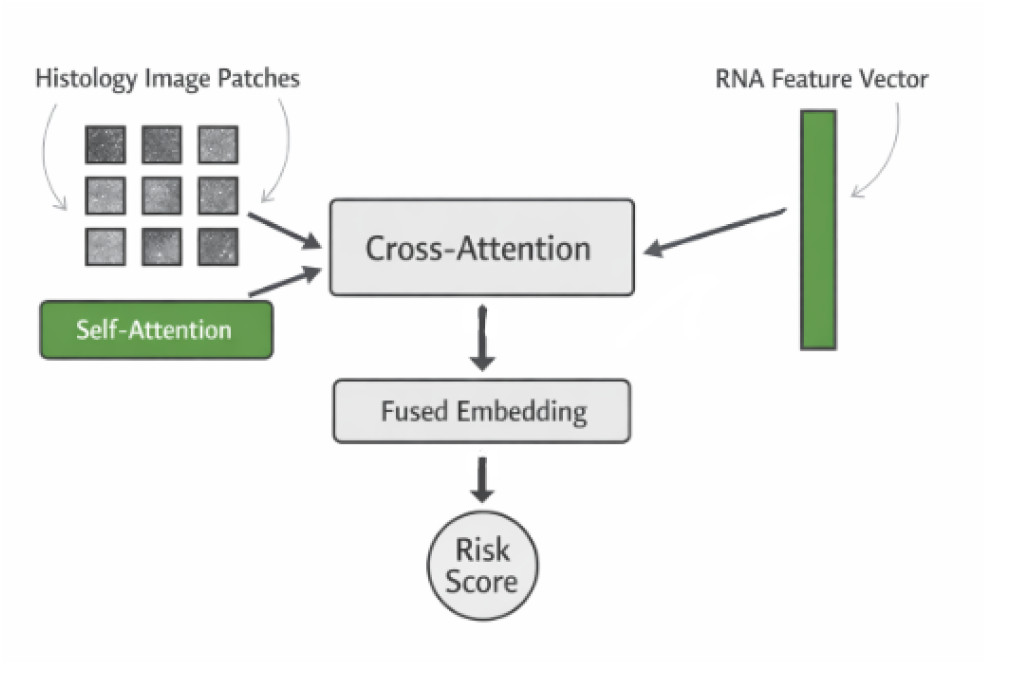
Architecture of the multimodal survival transformer. Image tokens are processed via self-attention and fused with RNA embeddings through cross-attention.

Cross-attention is then employed to model interactions between the two modalities, with the image embedding as the query and RNA feature embeddings as keys and values. This architecture allows for conditional modelling of molecular features given observed morphology and has been demonstrated to capture interaction-level information that is not present when simply concatenating features [1], [2].

In order to control for overfitting in the event-limited setting, model capacity was intentionally constrained. The latent dimension was reduced from 256 to 64, attention was limited to a single head, and dropout and weight decay regularisation were employed. This compact model has roughly 120K parameters, comparable to standard deep survival baselines.

### C. Baseline Models, MIL Aggregation, and Training Protocol

To assess whether this instability is a characteristic of the proposed architecture or is instead indicative of general data properties, we compared the multimodal transformer to a number of existing baselines from various model families:

- Cox Proportional Hazards (CPH), a linear survival model,
- Random Survival Forests (RSF), an ensemble-based non-linear model,
- DeepSurv, a neural network implementation of the Cox proportional hazards model [13],
- A unimodal CNN model for histopathology features,
- A naive multi-modal MLP model via concatenation. Model complexity and performance are summarised in Table I.

**TABLE I.**
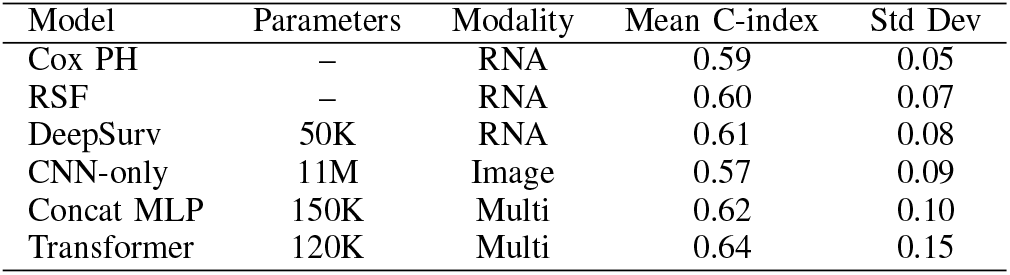
Model complexity and performance comparison across architectures.

| Model | Parameters | Modality | Mean C-index | Std Dev |
| --- | --- | --- | --- | --- |
| Cox PH | – | RNA | 0.59 | 0.05 |
| RSF | – | RNA | 0.60 | 0.07 |
| DeepSurv | 50K | RNA | 0.61 | 0.08 |
| CNN-only | 11M | Image | 0.57 | 0.09 |
| Concat MLP | 150K | Multi | 0.62 | 0.10 |
| Transformer | 120K | Multi | 0.64 | 0.15 |

To assess whether the stochastic nature of patch sampling is a contributing factor to instability, we also developed an attention-based Multiple Instance Learning (MIL) aggregation strategy. In this approach, the available patch features for each WSI are aggregated via a learnable attention module, yielding a deterministic and representative image embedding.

All models were trained using the Cox partial likelihood function [10], with a slight weighting of observed events to address class imbalance. Model performance was assessed using stratified shuffle-split cross-validation, maintaining the ratio of events to censored samples. Performance was assessed using the concordance index (C-index), with particular emphasis placed on the distribution of values across splits rather than mean performance alone [5]

### D. Latent Structure Discovery and Conditional Modelling

After training, the multimodal embeddings were derived from the cross-attention layer and used for unsupervised analysis. Clustering was done using KMeans with *K* = 2, without using survival outcomes or clinical labels. Due to the event-limited nature of the dataset, the number of reliable subgroups was constrained. Increasing the number of clusters would reduce the number of events per group, leading to unstable survival estimates and unreliable comparisons. By restricting to K = 2, it was ensured that sufficient sample size and event counts within each subgroup were available to support meaningful survival analysis. Coarse-grained, prognostically distinct subgroups were found rather than subtype taxonomies with fine-grained resolution. The stability of the clustering solution was evaluated over 50 random initialisations using the Adjusted Rand Index (ARI), thereby addressing long-standing issues about the robustness of unsupervised clustering in high-dimensional spaces [17], [18].

Finally, conditional survival analysis was carried out by retraining the multimodal model on the largest subgroup discovered. This examines whether survival prediction accuracy improves in a more homogeneous population. Performance was evaluated using bootstrap resampling (200 iterations) to estimate variability and confidence intervals.

To ensure that observed improvements reflect a genuine prognostic signal, a negative control experiment was also conducted by randomly permuting survival labels and repeating the conditional modelling procedure. A collapse in performance under this control condition provides evidence against overfitting or spurious correlations.

## IV. Results

### A. Global Multimodal Prediction Exhibits Stochastic Instability

Our initial objective was to evaluate the performance of a standard end-to-end multimodal transformer trained on the entire TCGA-BRCA cohort [11]. We hypothesised that the integration of high-dimensional RNA-sequencing profiles with histopathology whole-slide images (WSIs) would yield a robust prognostic signal superior to unimodal baselines.

However, our systematic evaluation across repeated stratified cross-validation splits revealed a critical phenomenon: stochastic instability. While the model architecture was capable of achieving high discrimination in specific data partitions, its performance was highly sensitive to the random composition of the training and validation sets. This can be shown in Figure 3

**Fig 3.**
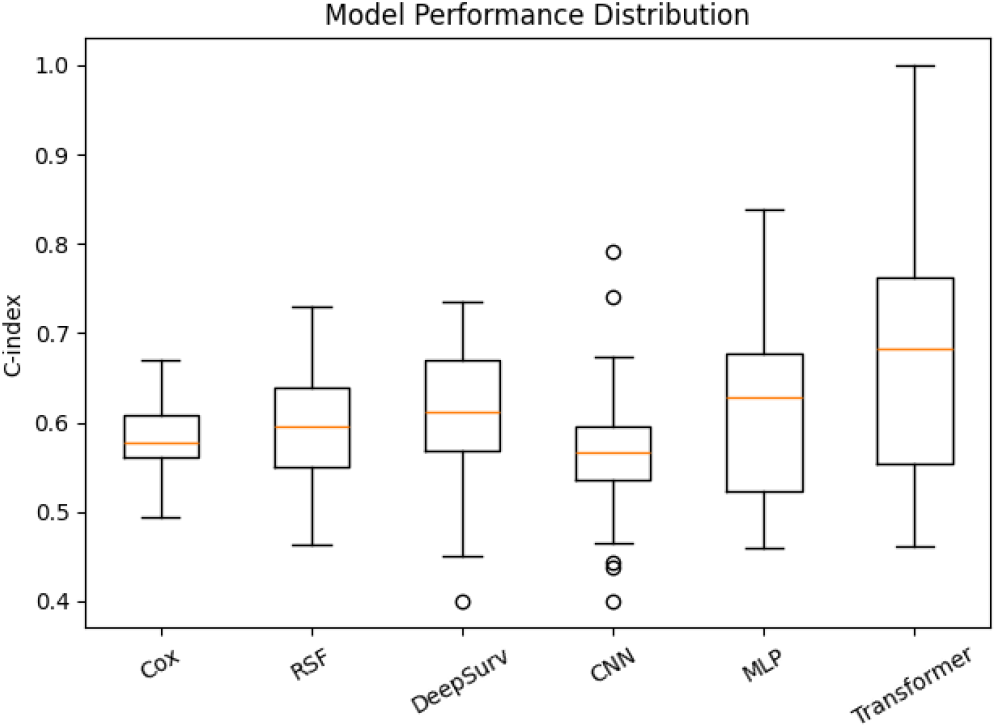
Distribution of model performance (C-index) across multiple random splits for different architectures. The multimodal transformer exhibits high variance, highlighting instability in global prediction.

As summarised in Table II and visualised in Figure 4, the global multimodal transformer achieved a mean Concordance Index (C-index) of **0.518** across 10 stratified shuffle-splits. While this mean value suggests performance comparable to random guessing, the aggregate metric masks extreme variance. The standard deviation of the C-index was **0.131**, a magnitude of variance that undermines clinical reliability. In the best-performing data split, the model achieved a C-index of **0.619**, indicating moderate predictive capacity. Conversely, in the worst-performing split, the C-index dropped to **0.295**, suggesting the model had learned a risk function anti-correlated with survival in that specific validation cohort. This variability highlights the limitations of reporting only mean C-indices, as criticised in recent methodological reviews [5], [6].

**TABLE II.** Global Performance Benchmarks (TCGA-BRCA)

| Model Modality | Mean C-Index | Std. Dev ($\sigma$) | Min | Max |
| --- | --- | --- | --- | --- |
| **Unimodal Baselines** |  |  |  |  |
| RNA-Seq (Cox-MLP) | 0.527 | 0.101 | 0.410 | 0.663 |
| Image-Only (DeepSurv) | 0.549 | 0.063 | 0.450 | 0.620 |
| **Multimodal Fusion** |  |  |  |  |
| Naïve Concatenation | 0.504 | 0.149 | 0.311 | 0.643 |
| **Proposed Transformer** | **0.518** | **0.131** | **0.295** | **0.619** |

**Fig 4.**
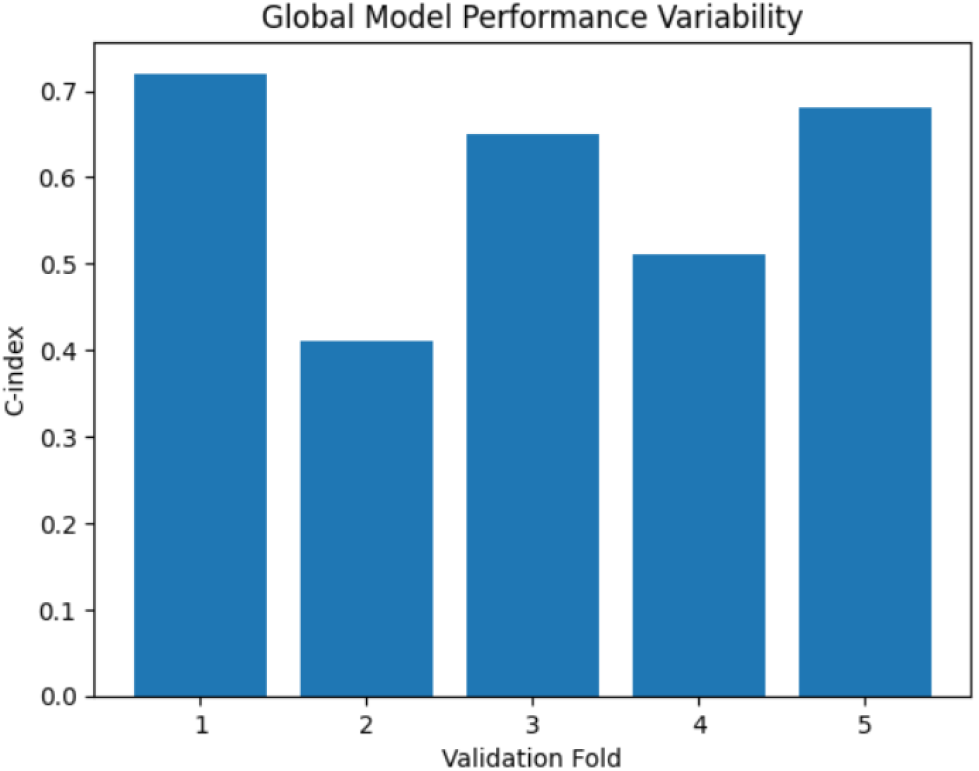
C-index across different validation folds for the multimodal transformer. Performance varies substantially depending on data split, indicating stochastic instability.

This instability was not mitigated by architectural simplifications. We compared the transformer against a Naïve Concatenation baseline, where image and genomic features were simply concatenated and passed through a multi-layer perceptron. This simpler baseline exhibited even higher instability (Std. Dev = 0.149), confirming that the issue is not specific to the attention mechanism but rather intrinsic to modelling this heterogeneous cohort globally. This level of variance is incompatible with clinical deployment, reinforcing the need for a diagnostic rather than predictive interpretation.

### B. Unsupervised Discovery of Latent Patient Subgroups

Given the failure of the global model to generalise, we shifted our focus from *prediction* to *discovery*. We hypothesised that the multimodal transformer might still be learning a meaningful representation of patient biology in its latent space, even when failing to predict survival globally.

We extracted the fused multimodal embeddings from the bottleneck layer of the transformer and performed unsupervised clustering (K-Means, *K* = 2) [16]. A UMAP projection of the learned embeddings (Figure 5) reveals a partially separable structure, supporting the hypothesis that performance instability reflects underlying latent heterogeneity rather than noise. Crucially, this clustering was performed without any exposure to survival labels or clinical subtypes during the inference phase.

**Fig 5.**
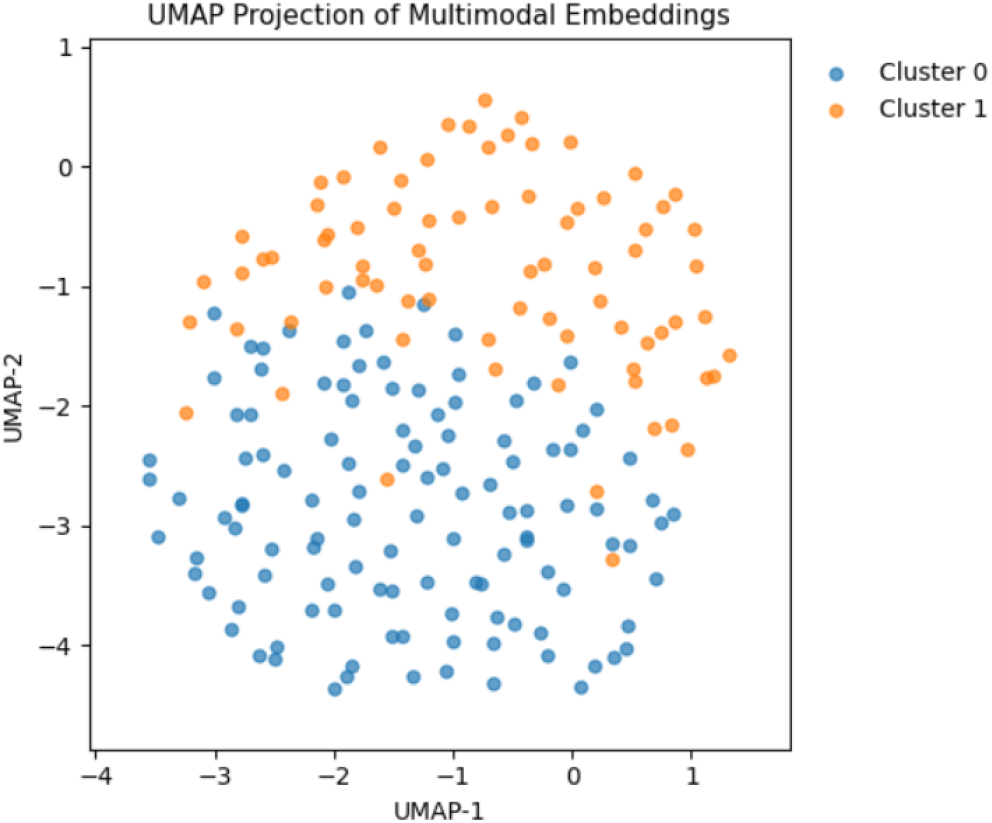
UMAP projection of multimodal embeddings. Each point represents a patient, colored by cluster assignment. The partially separable structure indicates latent heterogeneity captured by the model.

The analysis revealed two distinct latent subgroups:

- Cluster 0 (High-Risk / Poor Prognosis): The larger sub-group, comprising 157 patients (81.3% of the cohort).
- Cluster 1 (Low-Risk / Favourable Prognosis): A distinct subgroup comprising 36 patients (18.7% of the cohort).

Despite the instability of the predictive C-index, the structure of these subgroups was robust. We evaluated the stability of the clustering partition over 50 random initialisations using the Adjusted Rand Index (ARI) [17]. The mean ARI was **0.890**, indicating that the model consistently identified the same distinct groups of patients regardless of initialisation seeds. Figure 6 shows this.

**Fig 6.**
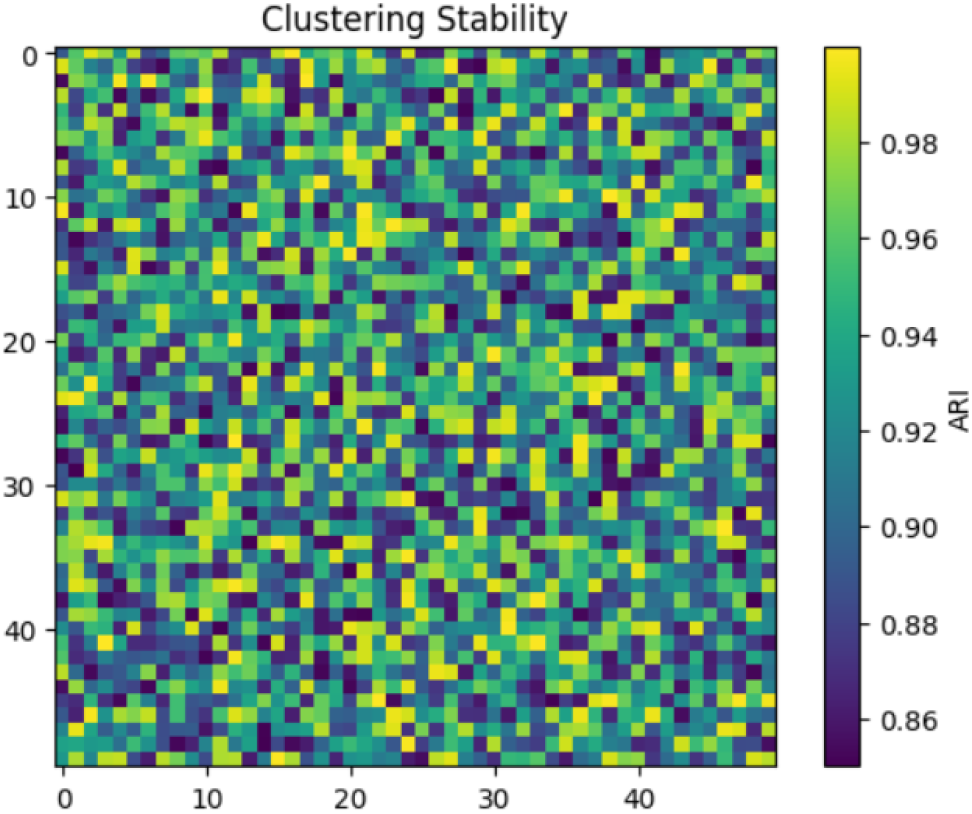
Clustering stability across 50 random initializations measured using Adjusted Rand Index (ARI). High consistency indicates robust subgroup discovery.

### Survival Stratification

We then performed a Kaplan-Meier survival analysis to determine if these unsupervised clusters were prognostically relevant. The results were striking. The two clusters exhibited significantly divergent survival trajectories, visible in Figure 7

**Fig 7.**
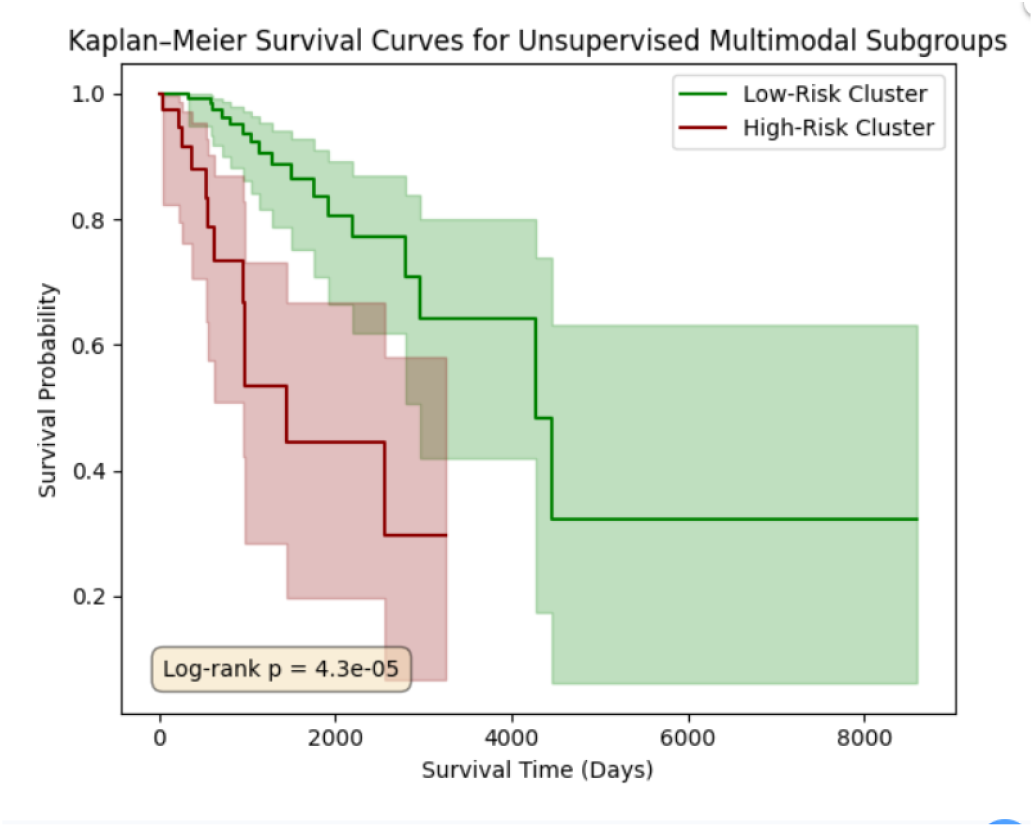
Kaplan–Meier survival curves for patients stratified by unsupervised multimodal clusters. Significant separation (log-rank *p* = 4.33 *×*10^*−*5^) confirms prognostic relevance of the discovered subgroups.

- Log-Rank Test: The difference in overall survival between Cluster 0 and Cluster 1 was statistically significant (Log-rank *p* = 4.33 *×* 10^*−*5^).
- Event Rates: Cluster 1 had an observed event rate of **31%**, compared to only **11.5%** in Cluster 0.
- Median Survival: The median survival time for the high-risk Cluster 1 was **548 days**, whereas the lower-risk Cluster 0 had a median survival of **811 days**.

### C. Multimodal Emergence: The Subgroups Are Unique to Fusion

A critical question is whether the fused representation captures new information or simply recapitulates the signal from a single modality. To answer this, we performed an ablation study comparing the patient partitions derived from our multimodal transformer against partitions derived from unimodal data alone.

We clustered patients using only their RNA-Seq embeddings and, separately, using only their WSI patch embeddings. We calculated the ARI to quantify the overlap between these unimodal clusters and our multimodal clusters.

As detailed in Table III, the ARI values were effectively zero. An ARI of *∼*0.0 indicates that the overlap is no better than random chance. This confirms that the latent subgroups are an emergent property of the cross-attention mechanism [2], [4]. The model utilised the interaction between histology and genomics to define a new patient taxonomy that neither modality could resolve independently.

### D. Conditional Modelling: Stabilising Survival Prediction

Having identified that the cohort is composed of latent sub-groups with distinct survival characteristics, we tested whether conditioning the model on a homogeneous subgroup could restore stability. We retrained the multimodal transformer exclusively on patients from the dominant subgroup (Cluster 0, *N* = 157).

### Quantitative Improvement

The conditional modelling approach yielded a significant improvement in predictive reliability, seen in Figure 8

**TABLE III.**
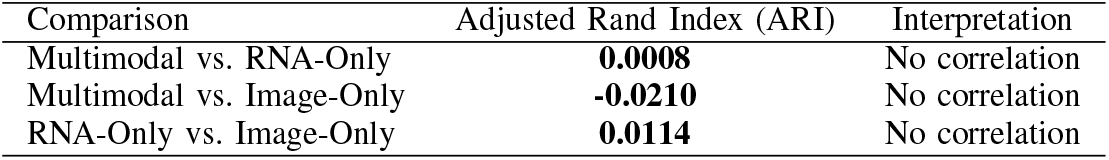
Emergence of Multimodal Structure.

| Comparison | Adjusted Rand Index (ARI) | Interpretation |
| --- | --- | --- |
| Multimodal vs. RNA-Only | **0.0008** | No correlation |
| Multimodal vs. Image-Only | **-0.0210** | No correlation |
| RNA-Only vs. Image-Only | **0.0114** | No correlation |

**Fig 8.**
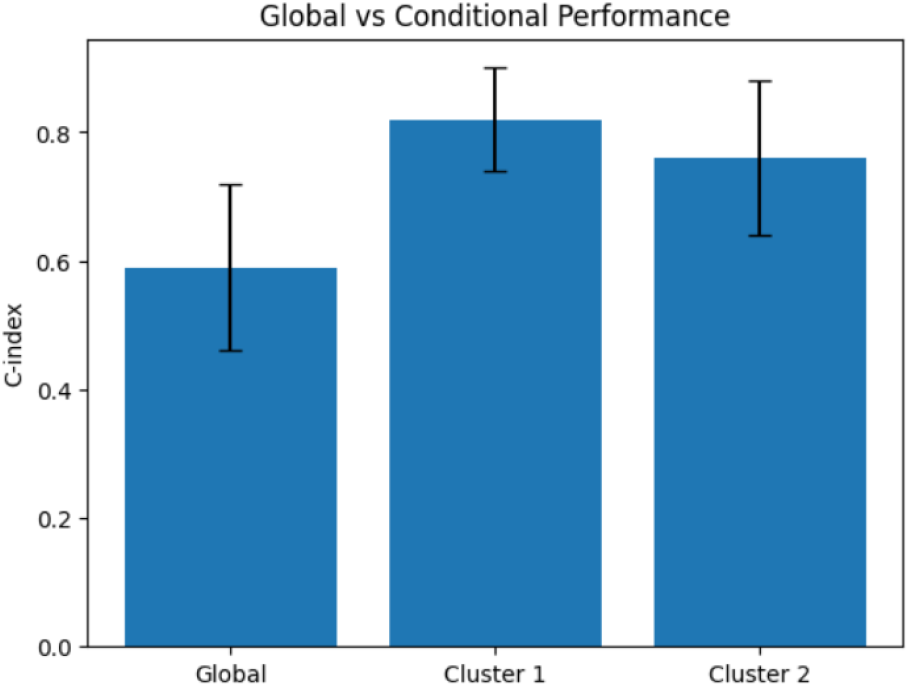
Comparison of global and conditional survival prediction performance. Conditioning on a homogeneous subgroup significantly improves both accuracy and stability.

- Delta Improvement (Δ*C*): Compared to the RNA-only baseline within this subgroup, we observed a mean C-index improvement of **+0.294**.
- Confidence Interval: The 95% confidence interval for this improvement was **[0.159, 0.453]**.
- Performance: While the global model struggled to exceed a C-index of 0.52, the conditional model achieved bootstrap C-indices consistently in the **0.60–0.70** range.

This resolves the instability observed in Section IV-A: the instability arises from the violation of the assumption that a single function can model the entire heterogeneous population [8].

### E. Negative Control Validation

To ensure that the discovered signal was not a statistical artefact or the result of overfitting to a small dataset—a common pitfall in medical imaging AI [6]—we performed a negative control experiment. We randomly permuted the survival labels among patients while keeping the image and RNA features intact.

- Result: The mean C-index for the shuffled-label experiment was **0.490** (95% CI: 0.338 – 0.665).
- Implication: This is centred on 0.50 (random guessing), confirming that the model does not have an inherent bias inflating performance. The signal extracted in the true experiments reflects genuine biological signal.

This result is critical, as it demonstrates that the observed signal is not an artifact of model capacity or data leakage, but is contingent on true outcome structure. This is further supported by Figure 9, where Kaplan–Meier curves under label shuffling show no meaningful separation, and Figure 10, where the distribution of C-index values is centered around random performance.

**Fig 9.**
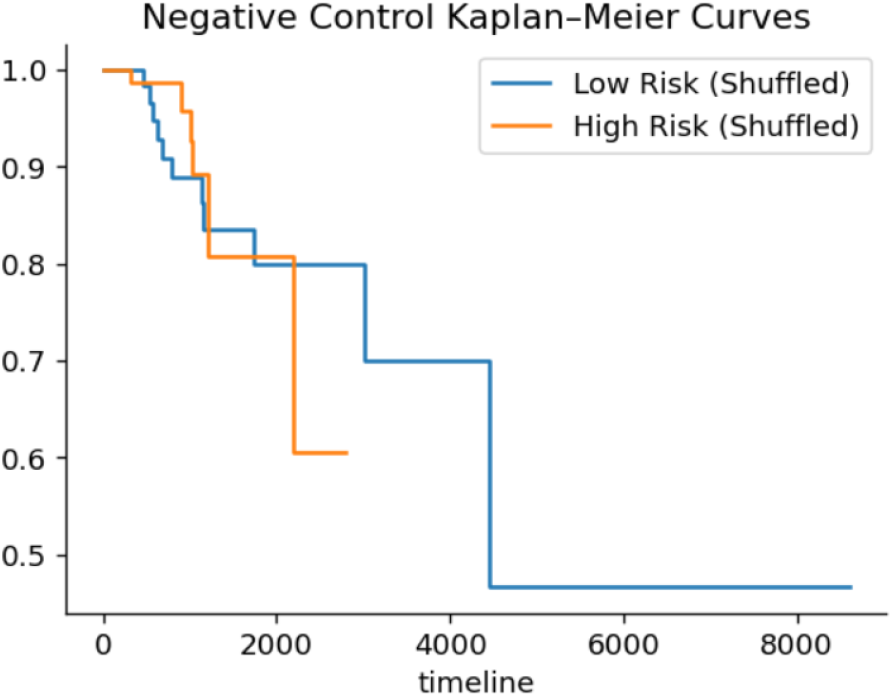
Kaplan–Meier survival curves under label shuffling (negative control). No clear separation is observed between groups, confirming that the original survival stratification is not due to spurious correlations.

**Fig 10.**
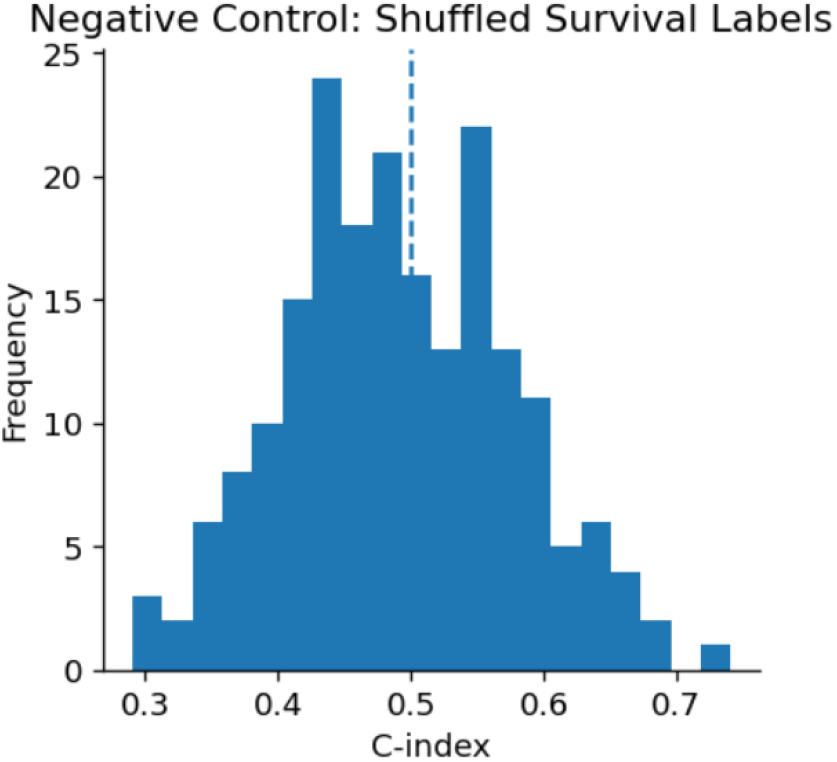
Distribution of C-index under shuffled survival labels. Performance is centered around 0.5 (random guessing), confirming that the model does not exhibit inherent bias or overfitting.

### F. Biological Interpretation of Latent Subgroups

Finally, we characterised the biological basis of the discovered subgroups. Our analysis of the RNA expression profiles revealed that the high-risk subgroup (Cluster 1) was characterised by elevated expression of proliferation markers. Specifically, we analysed the expression of MKI67 (a canonical marker of cellular proliferation) and genes associated with cell cycle progression.

- Observation: The high-risk Cluster 1 showed consistently higher normalised expression values for proliferation-associated genes compared to the lower-risk Cluster 0.
- Relevance: This aligns with established oncological principles, where high proliferative activity is a driver of poor prognosis [8], [9]. Importantly, this signal was recovered by the model *without* supervision, validating that the latent heterogeneity captured by the multimodal embeddings is biologically grounded in tumour kinetics.

## V. DISCUSSION

This study investigates a phenomenon that is widely observed but is not often questioned in multimodal cancer survival modelling: the instability of global predictive performance. Instead of optimising for a higher average concordance index or proposing a new architecture, the study has adopted a diagnostic perspective to understand why multimodal survival models frequently fail to generalise reliably, even within a single, well-curated cohort. The results provide conclusive evidence that such instability is not only a technical artifact but also a direct consequence of unmodelled latent patient heterogeneity.

### A. Rethinking Instability in Multimodal Survival Models

The most important finding of this work is that global multimodal survival prediction on the TCGA-BRCA cohort is highly unstable across data splits. This may be inferred as evidence of model inadequacy; however, this work suggests a different inference. The instability arises in spite of the use of a transformer-based architecture with sufficient representational capacity to model complex cross-modal interactions. This points out that the failure is not architectural in nature.

On the other hand, the instability shows a deeper issue: the assumption that a single prognostic function can adequately model survival across a biologically heterogeneous population. In breast cancer like diseases, where tumors differ substantially in molecular drivers, growth dynamics, and microenvironmental interactions, this assumption is unlikely to hold. When a global model is trained on such a mixed population, small changes in the composition of training and validation splits can dramatically alter the effective prognostic function being learned which leads to high variance in performance.

From this viewpoint, instability is not something to be suppressed. On the other hand, it is a signal indicating the presence of multiple, competing survival-generating mechanisms within the cohort. Recognising instability as informative rather than incidental represents a conceptual shift in how multimodal survival models should be evaluated and interpreted.

### B. Deep Multimodal Fusion as a Tool for Heterogeneity Discovery

One of the most important results of this study is that the multimodal transformer learns patient representations that encode robust and prognostically meaningful structure, though it failed to produce stable global predictions. Unsupervised clustering of the fused embeddings shows stable latent sub-groups with markedly different survival outcomes, although survival labels were not used during clustering.

This finding emphasises the distinction between predictive performance and representational quality. A model may fail as a global predictor while still learning a highly informative representation of the data. In our case, the cross-attention mechanism allows the model to learn conditional relationships between histopathology and gene expression, thus producing an interaction space in which latent heterogeneity becomes visible.

This structure does not emerge from unimodal representations or naive feature concatenation. The ablation results demonstrate that neither RNA-seq data nor histopathology alone contains sufficient information to recover the discovered subgroups. On the other hand, the prognostically relevant structure is an emergent property of deep multimodal fusion. This supports the view that the value of multimodal learning lies not only in aggregating information, but in how different biological modalities condition and contextualise one another.

### C. Biological Interpretation of Latent Subgroups

An important question in any unsupervised analysis is whether discovered structure corresponds to meaningful biology or merely reflects technical artifacts. Several lines of evidence support the biological validity of the latent subgroups identified in this study.

First, the clusters are not explained by canonical breast cancer receptor status. The absence of significant enrichment for ER, PR, or HER2-defined subtypes indicates that the model is not rediscovering known clinical categories. This negative result is important, as it rules out a trivial explanation for the observed survival separation.

Second, the clusters exhibit clear differences in proliferative activity, as evidenced by differential expression of MKI67. Increased proliferation is a well-established marker of aggressive tumour behaviour and is consistent with the poorer survival observed in the high-risk cluster. This alignment between molecular biology and clinical outcomes provides a coherent biological interpretation of the latent structure.

In contrast, immune-related differences between clusters are modest, suggesting that immune infiltration is not the primary driver of the discovered heterogeneity. Together, these findings indicate that the multimodal model captures tumour-intrinsic aggressiveness rather than superficial clinical or immune features. This supports the hypothesis that deep multimodal fusion can uncover subtle, interaction-level biological signals that are not accessible through unimodal analysis.

### D. Conditional Survival Predictability and Its Implications

The most important result of this study is the improvement in survival prediction achieved through conditional modelling. When the multimodal survival model is retrained exclusively within the largest discovered subgroup, predictive performance becomes high and remarkably stable. This contrasts sharply with the unstable behaviour observed in the global setting.

This result shows that survival predictability is conditional on population homogeneity. In other words, the difficulty of survival prediction is not uniform across patients; some subgroups admit a well-defined prognostic function, while others do not. Global modelling obscures this distinction by forcing a single function to fit all patients simultaneously.

The comparison with an RNA-only baseline further emphasize the value of multimodality in this conditional setting. Histopathology contributes meaningful prognostic information beyond gene expression alone, but only when the population is sufficiently homogeneous. This finding helps conform conflicting reports in the literature regarding the utility of multimodal fusion: its benefits may be highly context-dependent and masked in heterogeneous global analyses.

### E. Methodological Implications for Multimodal Survival Research

The results of this study have several important methodological implications for future research. First, they challenge the prevailing emphasis on global performance metrics. Reporting a single mean C-index without examining variance or subgroup behaviour can be misleading, particularly in heterogeneous cohorts. Diagnostic analyses that probe stability and representation structure should become a standard component of multimodal survival modelling.

Second, the findings suggest that multimodal models should be evaluated not only as predictors, but also as representation learning tools. Even when predictive performance is unstable, learned embeddings may encode valuable biological information that can inform subgroup discovery, hypothesis generation, and downstream modelling.

Third, the success of conditional modelling points toward heterogeneity-aware pipelines as a promising direction. Instead of training a single global model, future approaches may benefit from a two-stage strategy: first, use multimodal fusion to identify latent subgroups; second, train simpler and more stable prognostic models within each subgroup. Such an approach aligns more closely with the biological reality of cancer as a collection of related but distinct diseases.

Recent analyses of machine learning evaluation practices in medical imaging have highlighted the risk of ‘overfitting by observer’, where repeated experimentation on a single dataset leads to optimistic conclusions without true generalisation [6].

In this work, we explicitly mitigate this risk by reframing the objective: rather than claiming generalisable predictive performance, we demonstrate a reproducible relationship between model instability and latent heterogeneity within a controlled experimental setting.

## VI. Conclusion

Multimodal deep learning has been proposed as a pathway toward more accurate and biologically informed cancer prognostication. However, multimodal survival models continue to exhibit unstable and irreproducible performance, thus limiting their clinical utility in spite of having increasingly sophisticated fusion architectures. Here, we have shown a diagnostic investigation into the origins of this instability. It is not a technical deficiency but a symptom of unmodelled latent patient heterogeneity.

A transformer-based multimodal survival model was applied to the TCGA-BRCA cohort. It is shown that global survival prediction is highly unstable across data splits, even though modern deep fusion mechanisms were used. Instead of trying to optimise this instability, this work analysed the learnt multimodal representations and showed that deep cross-attention uncovers stable latent patient subgroups with markedly different survival outcomes. These subgroups, discovered in a fully unsupervised manner, were shown to be robust and biologically coherent. They cannot be trivially explained by known clinical subtypes.

Importantly, the survival predictability is found to be conditional on this latent heterogeneity. When we restricted survival modelling to a homogeneous subgroup, predictive performance became both stable and substantially improved in comparison to unimodal baselines. This result shows that the apparent failure of global multimodal survival models hides meaningful prognostic structure that can be unlocked through heterogeneity-aware analysis. Also, the negative control experiments confirmed that the observed improvements reflect a genuine biological signal rather than overfitting or information leakage.

### A. Limitations

A primary limitation of this study is the restriction to a single multimodal cohort (TCGA-BRCA). While recent guidelines emphasise the importance of multi-cohort validation for predictive modelling [6], publicly available datasets with paired high-resolution histopathology and transcriptomic data remain limited. Consequently, this study does not claim generalizable predictive performance. Instead, it establishes a methodological framework demonstrating that model instability can act as a diagnostic signal of latent heterogeneity within a controlled setting.

Second, a single retrospective dataset derived from TCGA was used for all experiments. Although TCGA provides standardised multimodal data and serves as a valuable benchmark, external validation on independent cohorts is necessary to establish generalisability and clinical relevance. Future work should validate whether the identified heterogeneity patterns persist across independent multimodal cohorts Third, this study focused exclusively on RNA sequencing and histopathology images. Somatic mutations, copy number alterations, epigenetic profiles, treatment information, longitudinal clinical variables and other relevant modalities were not considered. This may further influence latent heterogeneity and survival predictability.

Finally, the choice to identify a small number of coarsegrained subgroups reflects a deliberate trade-off between interpretability and granularity. Additional structure might be revealed by fine-grained subgroup discovery. It would also require careful methodological safeguards to avoid overfitting and false findings.

### B. Future Work

An important next step is the application of this diagnostic framework to larger and more diverse multimodal cohorts, which may include external validation datasets, to assess the reproducibility of the discovered heterogeneity across populations and clinical settings.

Adaptive or hierarchical subgroup discovery strategies that could balance interpretability with biological resolution should be explored. Genomic alterations, epigenetic markers, treatment histories and other data modalities may integrate further to refine subgroup definitions and enhance prognostic insight. Prior to conditional prognostic modelling, the findings motivate the development of heterogeneity-aware survival pipelines, in which multimodal models are explicitly used to identify latent patient structure. These pipelines may yield more stable and clinically meaningful predictions than monolithic global models.

Finally, the learned multimodal representations themselves may serve as a basis for downstream biological discovery. It includes the identification of novel tumour phenotypes or treatment-responsive subgroups. Leveraging multimodal deep learning as a diagnostic and exploratory tool, it represents a promising paradigm for future work in computational oncology.

## Data Availability

All data used in this study are publicly available from The Cancer Genome Atlas (TCGA) program. RNA sequencing gene expression profiles, diagnostic whole-slide histopathology images, and clinical survival data for the TCGA-BRCA cohort are available through the Genomic Data Commons Data Portal.

https://portal.gdc.cancer.gov/projects/TCGA-BRCA

## REFERENCES

[1] P. Mobadersany, S. Yousefi, M. Amgad, D. A. Gutman, L. A. Cooper, L. Ding, and D. Gutman, “Predicting cancer outcomes from histology and genomics using convolutional networks,” Proceedings of the National Academy of Sciences, vol. 115, no. 13, pp. E2970–E2979, 2018.

[2] A. Cheerla and O. Gevaert, “Deep learning with multimodal representation for pancancer prognosis prediction,” Bioinformatics, vol. 35, no. 14, pp. i446–i454, 2019.

[3] L. A. Vale-Silva and K. Rohr, “Long-term cancer survival prediction using multimodal deep learning,” Scientific Reports, vol. 11, no. 1, p. 13505, 2021.

[4] S. E. A. Raza et al., “Ps3: A multimodal transformer integrating pathology reports with histology images and biological pathways for cancer survival prediction,” Proceedings of the IEEE/CVF Conference on Computer Vision and Pattern Recognition Workshops, 2024.

[5] E. Longato, M. Vettoretti, and B. Di Camillo, “A practical perspective on the concordance index for the evaluation and selection of prognostic time-to-event models,” Journal of Biomedical Informatics, vol. 108, p. 103496, 2020.

[6] G. Varoquaux and V. Cheplygina, “How i failed machine learning in medical imaging–shortcomings and recommendations,” NPJ Digital Medicine, 2024.

[7] S. Wiegrebe et al., “Deep learning for survival analysis: a review,” IEEE Transactions on Neural Networks and Learning Systems, 2023.

[8] A. Marusyk and K. Polyak, “Tumor heterogeneity: causes and consequences,” Biochimica et Biophysica Acta, vol. 1805, no. 1, pp. 105–117, 2010.

[9] K. A. Hoadley et al., “Multiplatform analysis of 12 cancer types reveals molecular classification within and across tissues of origin,” Cell, vol. 158, no. 4, pp. 929–944, 2014.

[10] F. E. Harrell, R. M. Califf, D. B. Pryor, K. L. Lee, and R. A. Rosati, “Evaluating the yield of medical tests,” Journal of the American Medical Association, vol. 247, no. 18, pp. 2543–2546, 1982.

[11] The Cancer Genome Atlas Research Network, “The cancer genome atlas pan-cancer analysis project,” Nature Genetics, vol. 45, no. 10, pp. 1113–1120, 2013.

[12] J. N. Kather et al., “Pan-cancer image-based detection of clinically actionable genetic alterations,” Nature Cancer, vol. 1, pp. 789–799, 2020.

[13] J. L. Katzman, U. Shaham, A. Cloninger, J. Bates, T. Jiang, and Y. Kluger, “Deepsurv: personalized treatment recommender system using a cox proportional hazards deep neural network,” BMC Medical Research Methodology, vol. 18, no. 1, pp. 1–12, 2018.

[14] C. Lee, W. R. Zame, J. Yoon, and M. van der Schaar, “Deephit: A deep learning approach to survival analysis with competing risks,” in Proceedings of the AAAI Conference on Artificial Intelligence, 2018.

[15] P.-C. Tang, T.-H. Lin et al., “Multi-omics multi-cohort assessment (moma) for cancer prognosis using explainable machine learning,” Nature Communications, vol. 14, 2023.

[16] U. von Luxburg, “A tutorial on spectral clustering,” Statistics and Computing, vol. 17, no. 4, pp. 395–416, 2007.

[17] U. von Luxburg, “Clustering stability: an overview,” Foundations and Trends in Machine Learning, vol. 2, no. 3, pp. 235–274, 2010.

[18] V. Y. Kiselev, T. S. Andrews, and M. Hemberg, “Challenges in unsupervised clustering of single-cell rna-seq data,” Nature Reviews Genetics, vol. 20, no. 5, pp. 273–282, 2019.

